# Public perception of ethical issues related to COVID-19 control measures in Singapore, Hong Kong, and Malaysia: A cross-sectional survey

**DOI:** 10.1101/2021.03.01.21252710

**Authors:** Teck Chuan Voo, Angela Ballantyne, Ng Chirk Jenn, Benjamin J. Cowling, Jingyi Xiao, Phang Kean Chang, Sharon Kaur, Grazele Jenarun, Vishakha Kumar, Jane Mingjie Lim, Zaw Myo Tun, Nigel Chong Boon Wong, Clarence C. Tam

## Abstract

**Background:** Several countries have implemented control measures to limit SARS-CoV-2 spread, including digital contact tracing, digital monitoring of quarantined individuals and testing of travelers. These raise ethical issues around privacy, personal freedoms and equity. However, little is known regarding public acceptability of these measures.

**Methods:** In December 2020, we conducted surveys among 3635 respondents in Singapore, Hong Kong and Malaysia to understand public perceptions on the ethical acceptability of COVID-19 control measures.

**Findings:** Hong Kong respondents were much less supportive of digital contact tracing and monitoring devices than those in Malaysia and Singapore. Around three-quarters of Hong Kong respondents perceived digital contact tracing as an unreasonable restriction of individual freedom; <20% trusted that there were adequate local provisions preventing these data being used for other purposes. This was the opposite in Singapore, where nearly three-quarters of respondents agreed that there were adequate data protection rules locally. In contrast, only a minority of Hong Kong respondents viewed mandatory testing and vaccination for travelers as unreasonable infringements of privacy or freedom. Less than two-thirds of respondents in all territories were willing to be vaccinated against COVID-19, with a quarter of respondents undecided. However, support for differential travel restrictions for vaccinated and unvaccinated individuals was high in all settings.

**Interpretation:** Our findings highlight the importance of socio-political context in public perception of public health measures and emphasize the need to continually monitor public attitudes towards such measures to inform implementation and communication strategies.

**Funding:** This work was funded by the World Health Organization.

**Research in context:** *Evidence before this study:* We searched PubMed and Google Scholar for research articles published between 29 February 2020 to 20 January 2021 to identify empirical studies on public perception of restrictive and control measures imposed during COVID-19. We used the following terms: “COVID-19”, “SARS-COV-2”, “pandemic”, “public”, “population”, “survey”, “cross-sectional”, “national”, “international”, “perception”, “attitudes”, “opinions”, “views”, “acceptance”, “acceptability”, “support”, “ethics”, “restrictive measures”, “restrictions”, “control measures”, travel”, “contact tracing”, “testing”, “tests”, “quarantine”, “monitoring”, “vaccines” “vaccination”, “immunity”, “certificates”, “passports”, “digital”, “applications”, “apps”, “mandatory” and “compulsory”. We found 4 peer-reviewed publications: three population surveys on public acceptance of and ethical issues in digital contact tracing in France, Jordan, and Ireland, and one population survey on perceptions of immunity and vaccination certificates in Geneva, Switzerland. We found no studies that studied the relative acceptance of different types of control measures.

*Added value:* There is a paucity of literature on public perception of the ethics of control measures that have been or may be implemented in response to the COVID-19 pandemic. In this study, we found differing levels of public support in Singapore, Hong Kong, and Malaysia for digital contact tracing, wearable quarantine monitoring devices, and mandatory testing and vaccination for travelers. Hong Kong respondents sharply differed from Singapore and Malaysia respondents on perceptions of risks and benefits, the extent of intrusion into individual freedom, and assurance of privacy and data protection related to use of digital contact tracing and monitoring devices. These differences are likely to be substantially influenced by socio-political climate and governmental trust. Although less than two-thirds of respondents in all territories expressed a willingness to be vaccinated against COVID-19, we found high support for differential travel restrictions for vaccinated and unvaccinated individuals in all settings.

*Implications of all the available evidence:* Our survey provides evidence of strong public support of vaccination requirements for travelers within an Asian context, and differential restrictions for vaccinated and non-vaccinated travelers. It highlights the importance of wider socio-political influences on public perception and ethical issues related to control measures and emphasizes the need to continually monitor public attitudes towards such measures to inform implementation and communication strategies.

## Background

Several countries have implemented control measures to limit SARS-CoV-2 spread, including digital contact tracing, digital monitoring of quarantined individuals and testing of travelers. The rollout of COVID-19 vaccines means that vaccine acceptance, as well as vaccination requirements for travelers as a means to ease travel-related restrictions, are likely to be important considerations for policymakers.

Singapore, Hong Kong and Malaysia have all implemented policies in relation to digital contact tracing, monitoring of individuals during quarantine and testing of travelers. As of January 2021, COVID-19 testing and quarantine in hotels or designated quarantine facilities is mandatory for incoming travelers in all three territories. Digital contact tracing is implemented to different extents. The official digital contact tracing apps in Hong Kong (LeaveHomeSafe) and Malaysia (MySejahtera) can be used to record visits to premises by scanning a QR code on entry, report a positive COVID-19 test and receive notifications of potential exposure to COVID-19 cases. Although use of the app is voluntary, premises in Malaysia are required to record visits through the app. In Singapore, digital contact tracing comprises a combination of SafeEntry, which is used to record visits to premises, and TraceTogether, which uses Bluetooth technology either within a mobile phone app or standalone wearable token to record proximity to other users. Although TraceTogether is technically voluntary, its use is mandated for schoolchildren and for entry to certain venues.

These types of control measures raise ethical issues around restrictions on personal freedoms, invasion of privacy, and fairness or equity in the distribution of benefits and burdens.^1-6^ Despite this, relatively little is known regarding the public’s perceptions and acceptability of these measures.^7-10^ Acceptance is likely to depend on numerous factors, including the epidemic situation, perceived effectiveness, individual burdens, trust in authorities, and mechanisms to ensure data privacy. We conducted surveys in Singapore, Hong Kong and Malaysia to understand public perceptions on the ethical acceptability of digital contact tracing, wearable quarantine monitoring devices, and travel-related COVID-19 testing and vaccination measures.

## Methods

The surveys were conducted during December 2020 in all three territories. In Singapore and Hong Kong, respondents were recruited from population-representative online panels. In Malaysia, a market surveyor recruited respondents at shopping malls, community town halls and residential halls. Respondents were adults aged 18 years or older in Hong Kong and Malaysia, and 21 years or older in Singapore, corresponding to the minimum age of consent in each setting. Details of sampling and recruitment procedures are given in the Supplementary Information.

### Survey Questionnaire

We used the same questionnaire in all three territories, with modifications made to collect setting-specific socio-demographic information for comparison with census statistics. The questionnaire comprised five sections that probed general attitudes to vaccines, and opinions on digital contact tracing, use of wearable monitoring devices during quarantine, and COVID-19 testing and vaccination. We measured general vaccine attitudes using the Vaccine Confidence Scale.^11^ Respondents were also asked whether they would be willing to take a safe and effective COVID-19 vaccine when it became available.

To measure public opinion on the use of monitoring devices during quarantine and digital contact tracing, participants were asked to register their level of agreement with a series of statements using a 5-point scale. These statements pertained to perceived effectiveness and benefits of digital contact tracing, concerns about privacy and personal freedom, and trust in governance of contact tracing data.

Additionally, we asked respondents for their opinions on travel-related COVID-19 testing and vaccination policies. To measure public perception of policies related to COVID-19 vaccination for travel, we used two different scenarios: in the first, participants were asked to imagine that a COVID-19 vaccine had been approved for general use and is widely available, while in the second, participants were asked to imagine that a COVID-19 vaccine had been approved but was in limited supply. The core questions under each scenario were the same and the two scenarios were administered to randomly selected subsets of survey participants, to allow for comparison of whether vaccine availability influences people’s perception of vaccination policies. For instance, we asked participants if they thought it would be reasonable to allow travel only for vaccinated individuals or that additional restrictions be placed on unvaccinated travelers. Lastly, to measure vaccination intention, we asked participants if they would be willing to get a COVID-19 vaccine to travel abroad.

### Data analysis

We assessed representativeness of survey samples by comparing the socio-demographic characteristics of the survey sample from each site with those from their national census in terms of age group, gender, ethnicity, educational level, and socioeconomic status.

For each territory, we computed response frequencies and percentages for each survey question. For socio-demographic variables with notable deviations from the census distribution, we assessed the impact on survey responses by using post-stratification weights, to re-weight sample responses in proportion to the census distribution. For each respondent, we calculated scores for vaccine confidence, support for use of monitoring devices during quarantine, and support for digital contact tracing (Supplementary Information Tables S9-S11)

We further investigated whether willingness to be vaccinated against COVID-19 was influenced by general confidence in vaccines, and level of support for use of monitoring devices during quarantine and digital contact tracing using Spearman’s correlation coefficient.

All analyses were performed using R version 4.0.3.^12^

### Ethics Statement

Ethical approval for this study was obtained from the Ethics Review Committee of the Saw Swee Hock School of Public Health, National University of Singapore (SSHSPH-092); Universiti Malaya Research Ethics Committee (UM.TNC2/UMREC_1129); and the Institutional Review Board of the University of Hong Kong (UW-20-095).

## Results

There were 982 eligible respondents in Singapore, 1974 in Malaysia and 679 in Hong Kong. In Singapore, the survey sample was comparable to the census population in terms of marital status and housing type, but there was an over-representation of females and those with post-secondary and tertiary education, while those of Malay ethnicity and people in the highest income bracket were under-represented. In the Malaysia sample, those aged 30-49 years, females, those of Chinese ethnicity, unmarried individuals and those living in condominiums or single occupancy housing were overrepresented compared to the census population. In Hong Kong, the survey sample comprised proportionately more males, people with tertiary education, people in the highest income bracket and those living in public housing (Supplementary Information Tables S1-S3).

### Vaccine confidence and willingness to be vaccinated against COVID-19

In general, vaccine confidence was higher in Malaysia compared with Singapore and Hong Kong (Figure 1). Around two-thirds of respondents in Malaysia agreed that vaccines are safe, effective and an important health intervention for children. In addition, >75% agreed that vaccines are compatible with their religious beliefs. This figure was higher than in both Singapore and Hong Kong, where less than two-thirds and less than half of respondents respectively felt that vaccines were compatible with their religious beliefs. Further, confidence in vaccines was considerably lower among Hong Kong respondents, with about 4 in 10 agreeing that vaccines are safe and around half believing that vaccines are effective and an important health intervention for children.

**Figure 1:**
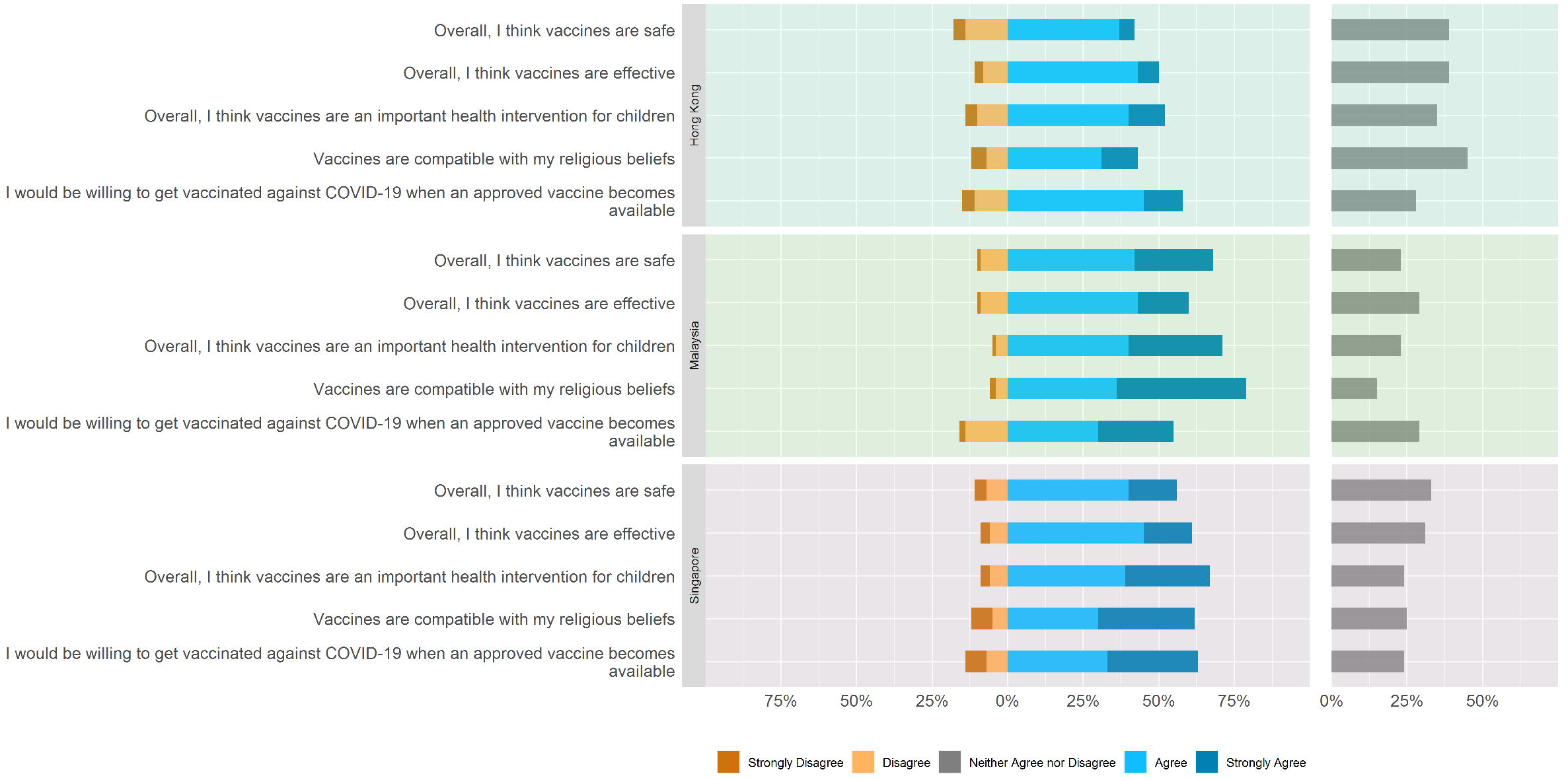
Vaccine confidence and willingness to be vaccinated against COVID-19 among respondents in Hong Kong, Malaysia and Singapore, December 2020

Across all three settings, two-thirds or fewer respondents said that they would be willing to be vaccinated against COVID-19 when an approved vaccine became available (Figure 1). Notably, about a quarter of respondents in all three settings were undecided about vaccination. Willingness to vaccinate was strongly associated with vaccine confidence in all three settings (Supplementary Information Tables S12-S14). Willingness to vaccinate was also higher among those displaying greater support for digital contact tracing and use of monitoring devices during quarantine (Supplementary Information Figures S1-S2).

### Concerns about invasiveness of different control measures

In general, respondents in Singapore and Malaysia demonstrated greater support for digital contact tracing technologies compared with Hong Kong. Around 75% of Singapore respondents believed that digital contact tracing was effective at reducing the risk of COVID-19 spread and around two-thirds felt that the benefits of digital contact tracing outweighed the harms. In contrast, only a quarter of Hong Kong respondents agreed that the benefits outweighed the risks and slightly more than a third believed such technologies to be effective at reducing risk of COVID-19 spread.

Differences were also seen between settings in the perceived intrusiveness of different control measures. Hong Kong respondents were much more likely than Singapore respondents to perceive that measures such as mandatory use of digital contact tracing technologies and monitoring devices during quarantine were unreasonable restrictions of individual freedom, with Malaysia respondents in between these two extremes (Figure 2). Interestingly, in Hong Kong, mandatory use of testing and vaccination for travelers were viewed far more positively; only a minority of respondents believed these measures to be unreasonable infringements of privacy or freedom. In Singapore, testing was considered less intrusive but vaccination more intrusive compared with digital contact tracing and use of monitoring devices. In Malaysia, intrusiveness concerns were similar for all measures, with around 50% of respondents stating that these were unreasonable intrusions.

**Figure 2:**
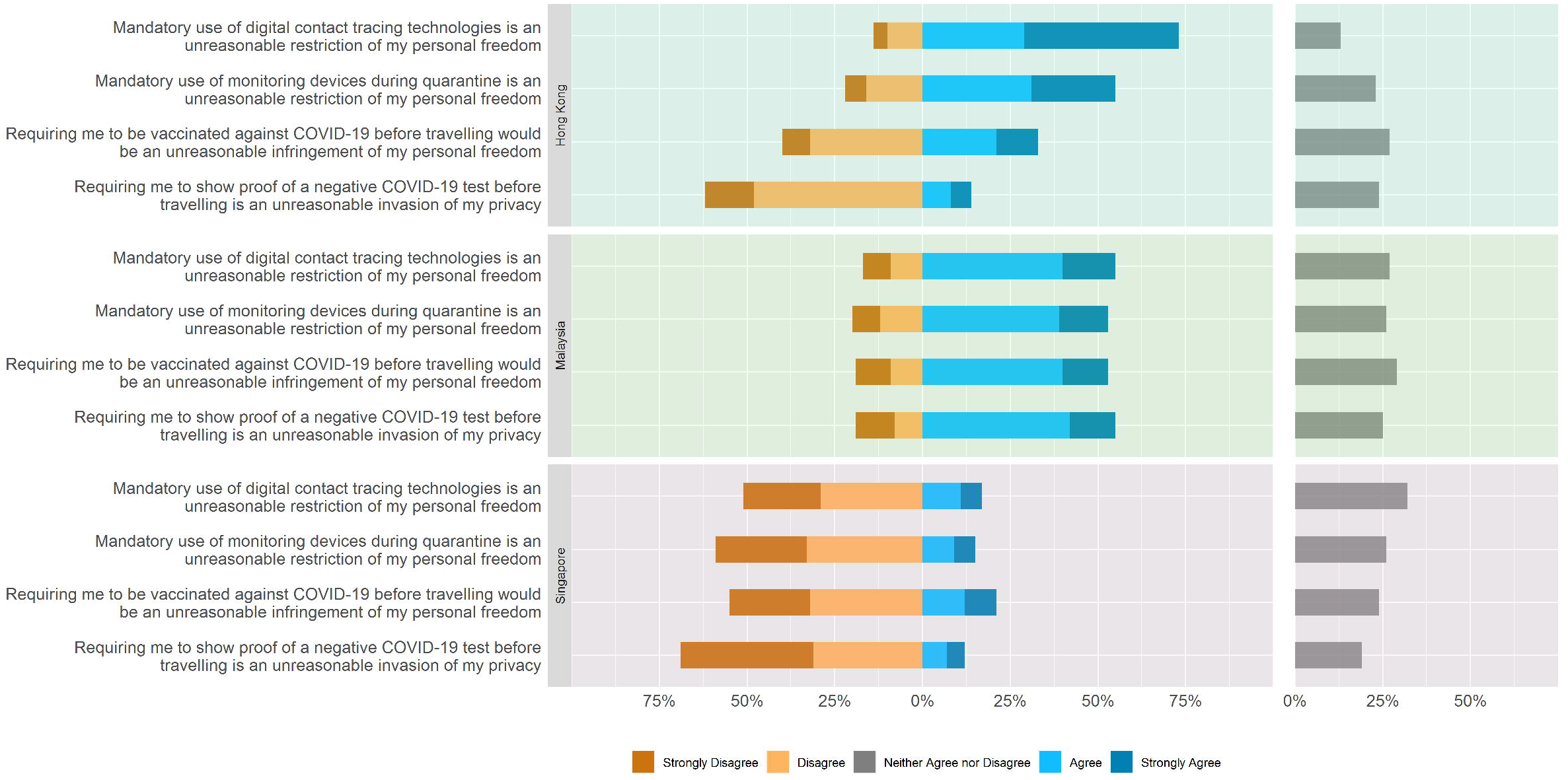
Perceived intrusiveness of different control measures among respondents in Hong Kong, Malaysia and Singapore, December 2020

These differing views were also reflected in differences in the level of trust in how digital contact tracing data would be used in different settings. In Hong Kong, <20% trusted that there were strict rules in place locally to prevent these data from being used for other purposes, while slightly over a third of respondents trusted that there were such rules in place in other countries. This was the opposite in Singapore, where nearly three-quarters of respondents agreed that there were adequate data protection rules in place locally, but less than a third felt that this was the case in other countries. In Malaysia, two-thirds of respondents agreed that there were adequate data protection rules in place both locally and in other countries (Figure 3). Similarly, support for mandatory use of digital contact tracing during the pandemic was low in Hong Kong. Only a minority agreed that this technology could help reduce the risk they posed to others if they became infected, or that digital contact tracing was a way for them to contribute to pandemic control efforts. In contrast, more than two-thirds and three-quarters of respondents in Malaysia and Singapore respectively agreed with these statements.

**Figure 3:**
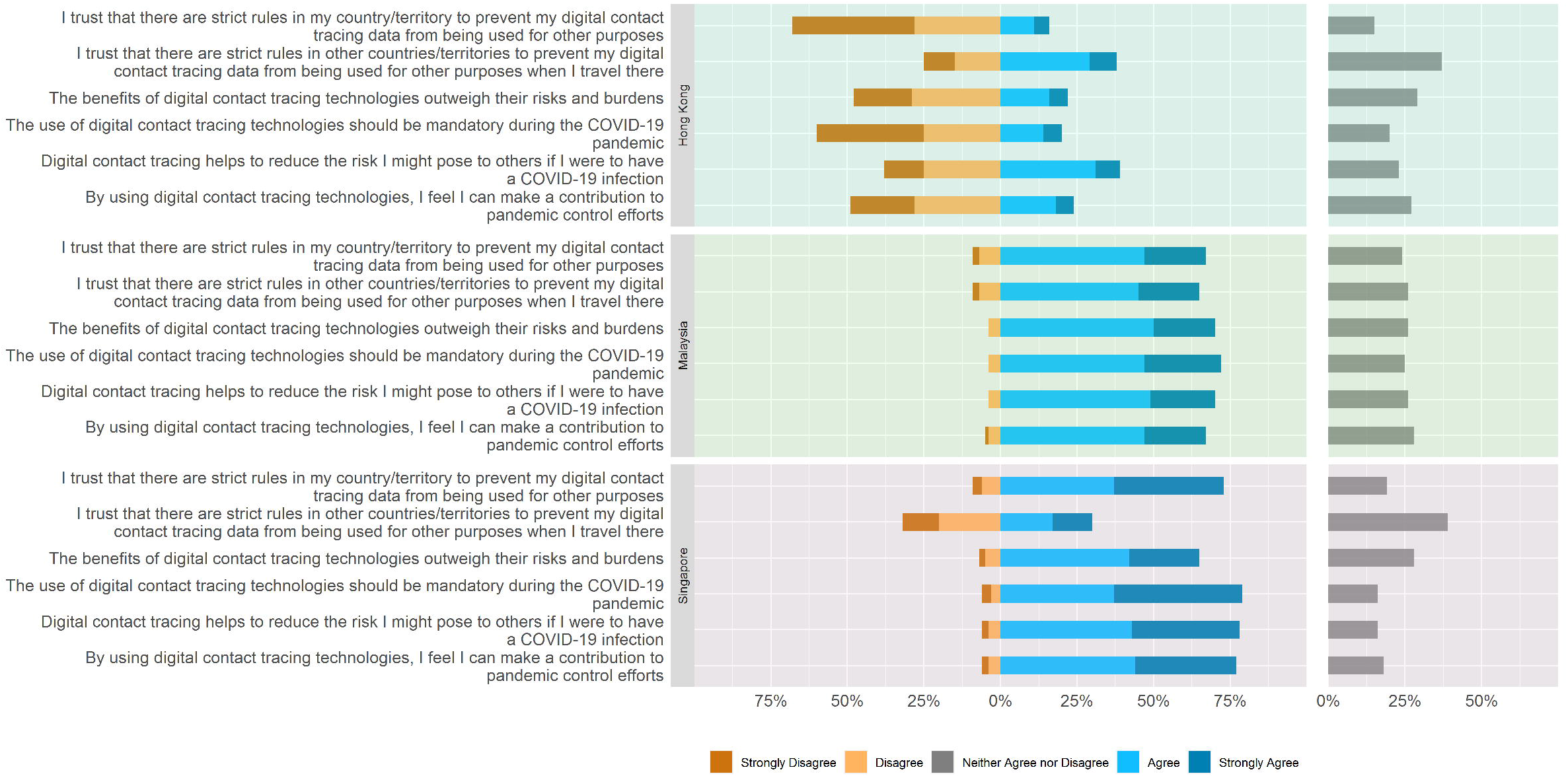
Digital contact tracing: privacy, trust, and mandatory use among respondents in Hong Kong, Malaysia and Singapore, December 2020

### Perceptions of vaccination policies for travelers

Support for travel-related COVID-19 vaccination was higher in Malaysia than in the other two settings; around two-thirds of respondents in Malaysia agreed that it was reasonable to allow travel only for vaccinated people, that it was reasonable to require all travelers to be vaccinated, and that it was reasonable to allow all travel for vaccinated people, but only essential travel for unvaccinated individuals. These figures were slightly lower in Singapore, while in Hong Kong less than half of respondents agreed with these statements (Figure 4).

**Figure 4:**
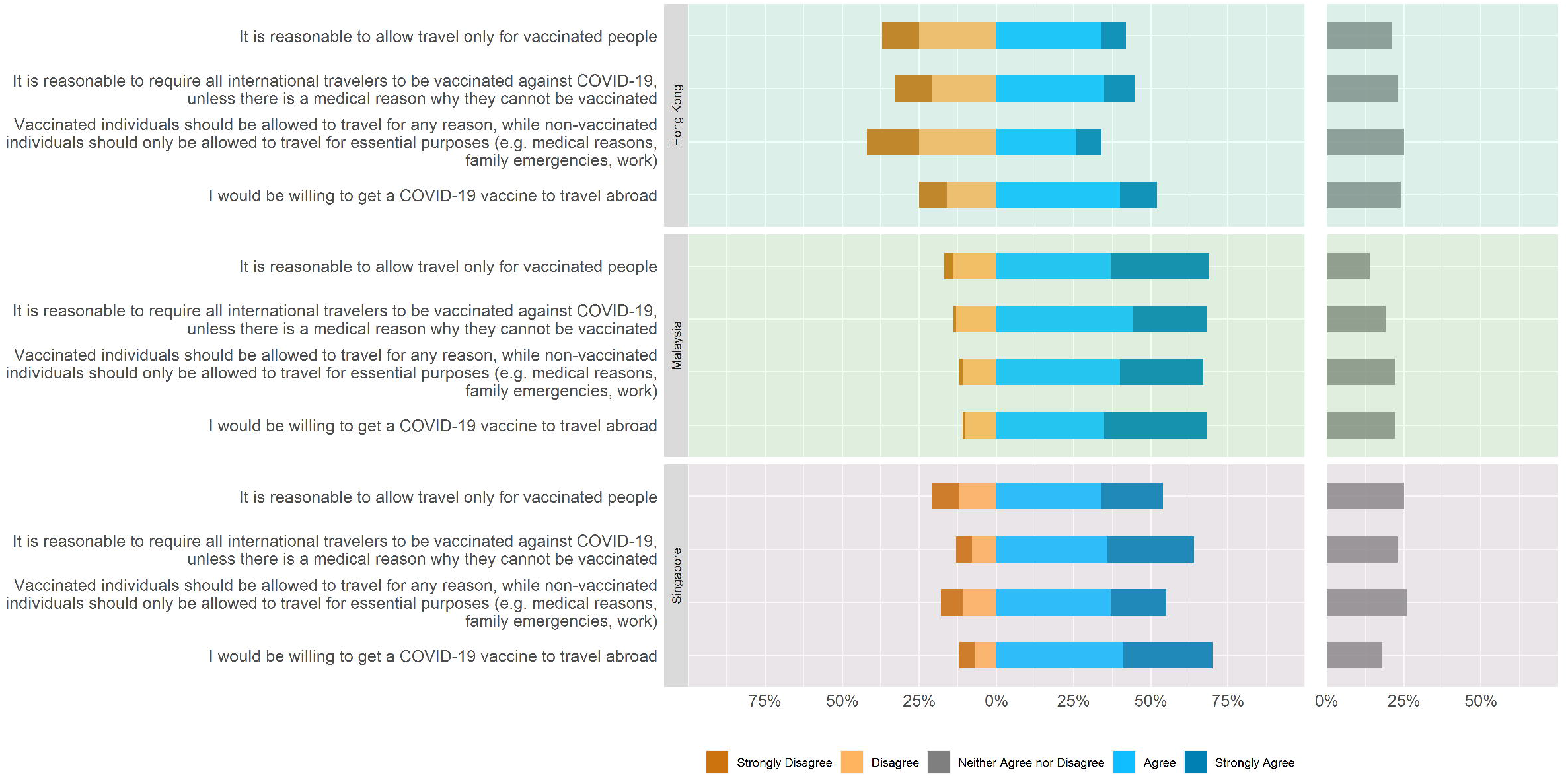
Perceptions of COVID-19 vaccination for travel among respondents in Hong Kong, Malaysia and Singapore, December 2020

Perceptions of equity implications of vaccination policies were mixed. Around a half of respondents in all three settings agreed that banning unvaccinated people from traveling internationally would be unfair. The majority of respondents in all three settings also agreed that it was reasonable to place different restrictions on vaccinated and unvaccinated travelers, and around a half agreed that it was reasonable for travelers to pay for vaccination, even if it meant that some groups in the population may not be able to afford to travel (Figure 5).

**Figure 5:**
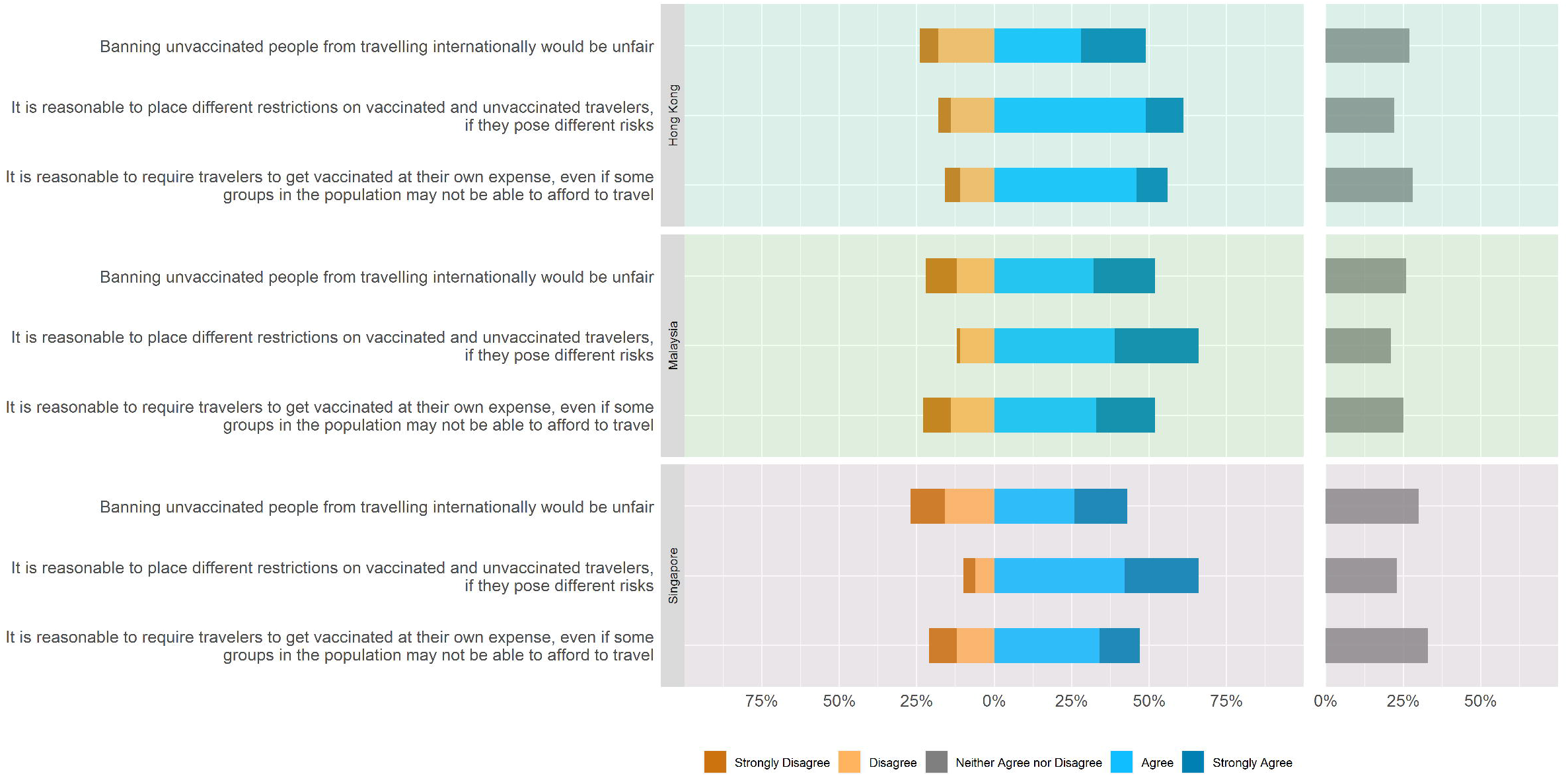
Equity considerations in travel-related COVID-19 vaccination among respondents in Hong Kong, Malaysia and Singapore, December 2020

Views on vaccination requirements for travel were not strongly influenced by whether vaccines were widely available or in limited supply, and applying weights to reflect the census distribution of demographic variables made little difference to the results (data not shown).

## Discussion

Like many countries and territories, Singapore, Malaysia and Hong Kong have adopted multiple strategies to ensure safe movement in community and international travel contexts during the COVID-19 pandemic. These include COVID-19 testing, wearable quarantine monitoring devices and digital contact tracing. The use of these control measures is likely to continue in many countries, at least during the pandemic, and could extend to vaccination certification to ease or lift movement and travel restrictions. The World Health Organization Emergency Committee on COVID-19 has recommended that countries should not introduce policies that require proof of vaccination from incoming travelers, given the limited global vaccine supply and uncertainty regarding whether vaccines reduce transmission risk.^13^ Nevertheless, the rationale for vaccination certification could change over time as scientific evidence accumulates and vaccine supply is ramped up. Understanding the public’s acceptance of these control measures is thus important for informing policy and communication strategies.

Justification for potentially intrusive control measures depends on a range of public health and ethical considerations such as effectiveness, necessity, proportionality, equitable distribution of benefits and burdens, and risk minimization.^14-15^ Acceptance, however, depends as much on the context as the intervention itself. Although on the face of it, both Hong Kong and Singapore have had similar success minimising community transmission of SARS-CoV-2 with similar measures focused on travel restrictions, strict quarantining, case isolation and testing, our study reveals stark differences in public opinion regarding the measures used to achieve this. Hong Kong residents display much lower support for surveillance and monitoring technologies such as digital contact tracing and wearable devices, lower trust in the governance of these technological measures, and greater opposition to their intrusiveness. Neighbouring Singapore and Malaysia, despite sharing many cultural similarities, also differ substantially in their views on the intrusiveness of control measures and implications for personal freedoms, including use of monitoring devices, digital contact tracing and testing for travel.

The ethics of surveillance, monitoring and health certification measures are context sensitive, contingent on a territory’s specific epidemic situation, inequalities, power structures and legal protections. Trust in these structures and protections is a fundamental element of social capital, especially in the context of a public health emergency, and an important determinant of citizens’ compliance with public health policies. Research during the 2014-2015 Ebola epidemic in Liberia found that people who distrusted their government took fewer precautions against Ebola and were less compliant with control measures,^16^ while a recent study in France showed that trust in government was highly associated with willingness to use digital contact tracing technology.^8^

During the 2002-2003 Severe Acute Respiratory Syndrome (SARS) epidemic, public trust in the Singaporean government to deal with the epidemic was shown to be high,^17^ while public trust in the Hong Kong government appeared to have been lower.^18^ The 2019 Edelman Trust Barometer also demonstrated higher trust in government (“do you trust the government to do what is right?”) in Singapore (67%) compared with Malaysia (60%) and Hong Kong (55%).^19^ Our research similarly demonstrates the influence of wider political and social conditions on people’s perception of public health interventions. For example, respondents from Hong Kong viewed contact tracing technology as less effective than respondents from Malaysia or Singapore. The effectiveness of the technology used for digital contact tracing in these three settings is unlikely to differ significantly. This difference in perceived effectiveness likely reflects a broader lack of trust in political institutions and data security in Hong Kong;^20-21^ more than two-thirds of respondents in Hong Kong expressed concern about domestic measures in place to prevent use of contact tracing data for other purposes.

Our study indicates that in Singapore, Hong Kong and Malaysia there is high public acceptance of COVID-19 testing to support safe international travel. Moreover, testing was considered less intrusive compared with use of monitoring devices during quarantine and digital contact tracing, particularly in Hong Kong. The reasons for this are unclear, but could be related to the lower burden of testing, perceived trust in how testing data are used, and individual benefit gained from knowing one’s test result.

Recent surveys have shown wide variability in the public’s willingness to be vaccinated against COVID-19, raising concerns about the potential impact of vaccine hesitancy on COVID-19 vaccination programs.^22-24^ Our survey indicates that willingness to vaccinate is modest in all three settings. Although we did not specifically ask about reasons why respondents were unwilling to be vaccinated, general confidence in vaccines was strongly related to willingness to be vaccinated against COVID-19. Importantly, a substantial fraction of the public in all three territories were undecided about whether to be vaccinated, which may indicate a need for more effective communication strategies to allay concerns among undecided individuals.

Nevertheless, support for travel-related vaccination was relatively high, with the majority of respondents in all three territories being in favor of vaccination requirements for travelers and showing willingness to be vaccinated for travel purposes. This is consistent with a recent population-based survey in Geneva, Switzerland, which showed general public support for vaccination requirements to allow foreign travel and strong support for policies that place different travel restrictions on vaccinated and unvaccinated individuals.^9^ From an ethical standpoint, allowing unvaccinated and vaccinated individuals to travel under different least restrictive conditions (consistent with minimizing public and individual health risks) would be equitable and respects individual freedom of movement.^25^

Our study has some potential limitations. For logistical reasons it was not possible to recruit Malaysian respondents through online panels as in the other two settings. The Malaysian sample was therefore younger relative to the general population, which may have affected the representativeness of opinions on control measures. However, re-weighting survey respondents to reflect the census distribution of demographic variables made no substantive difference to the results.

It should also be noted that public perceptions of control measures are dynamic and can change over time, particularly during an epidemic. In Singapore, our survey was conducted prior to the government’s announcement that digital contact tracing data could legally be used for criminal investigations, despite earlier public assurance that data would be used solely for contact tracing.^26^ In its efforts to maintain trust, the Singapore government passed a bill to limit criminal investigation uses of the data to specific serious offences, and set stronger penalties for data misuse than what is set out in current public sector data protection laws. In Hong Kong, the government recently announced that residents would be able to choose which vaccine to receive from those available in order to build trust in its vaccination program.^27^ Such policy developments influence the public discourse around public health control measures and likely shape the public’s opinion on the use of these measures over time. This emphasizes the need to continually monitor public perception of public health measures both during peacetime and during the course of a public health emergency. This can inform policy by providing an understanding of the limits of public acceptance of control measures in different contexts, identifying areas of concern to be addressed in the design, implementation and communication of control measures, and anticipating changes in public opinion that may affect acceptance or adherence to these measures. For example, a recommended intervention to promote public trust and acceptance of digital surveillance and monitoring is to implement an ethics oversight mechanism,^2, 28, 29^ but much will likely depend on how impartial, independent and inclusive this mechanism is regarded by the public. Routine community engagement and public deliberation exercises could be useful mechanisms to better understand and respond to specific local concerns, rather than relying on historical research, research findings from other settings, or on abstract normative analysis alone.

## Data Availability

All of the individual, de-identified participant data collected during the study as well as the study questionnaires will be available in our university's data warehouse or other public data repository beginning 9 months after article publication for an indefinite period.

## Acknowledgements

We thank the Singapore Population Health Studies (SPHS) unit for their assistance in pre-testing and data management for the questionnaire fielded to the online panel.

## Author contributions

Conceptualisation of study: VTC, CCT, AB; Development of study protocols: VTC, CCT, AJB, CJN, BJC, JX, PKC, SK, GJ, VK, JML, ZMT; Verification of data: NCBW, ZMT, CCT, JX, CJN Data analysis: NCBW, ZMT, CCT; Interpretation of data: CCT, ZMT, NCBW, VTC, AB, CJN, BJC, JX, PKC, SK, GJ; Manuscript drafting: VTC, CCT, AB; Editing of manuscript: All authors

## Data sharing statement

|  |  |
| --- | --- |
| Will individual participant data be available (including data dictionaries)? | Yes |
| What data in particular will be shared? | All of the individual, de-identified participant data collected during the study |
| What other documents will be available? | Study questionnaires |
| When will data be available? | Beginning 9 months after article publication for an indefinite period |
| With whom? | Anyone who wishes to access the data |
| For what types of analyses? | Any purpose |
| By what mechanism will the data be made available? | The data will be available in our university's data warehouse or other public data repository but without investigator support other than deposited metadata |

