## Supplementary Information for "Public perception of ethical issues related to COVID-19 control measures in Singapore, Hong Kong, and Malaysia: A cross-sectional survey"

Contributors:

Teck Chuan Voo,^a^ Angela Ballantyne,^b^ Ng Chirk Jenn,^c^ Benjamin J. Cowling,^d,e^ Jingyi Xiao,^ed^ Phang Kean Chang,^f^ Sharon Kaur,^g^ Grazele Jenarun,^h^ Vishakha Kumar,^i^ Jane Mingjie Lim,^i^ Zaw Myo Tun,^i^ Nigel Chong Boon Wong,^i^ Clarence C. Tam^i,j^

Affiliations

^a^ Centre for Biomedical Ethics, Yong Loo Lin School of Medicine, National University of Singapore, Singapore

^b^ Department of Primary Health Care and General Practice, University of Otago, Otago, New Zealand

^c^ Department of Primary Care Medicine, Faculty of Medicine, University of Malaya, Kuala Lumpur, Malaysia

^d^ WHO Collaborating Centre for Infectious Disease Epidemiology and ControlDivision of Epidemiology and Biostatistics, School of Public Health, The University of Hong Kong, Hong Kong Special Administrative Region, China

^e^ Laboratory of Data Discovery for Health Limited, Hong Kong Science and Technology Park, Hong Kong Special Administrative Region, ChinaLi Ka Shing Faculty of Medicine, The University of Hong Kong, Hong Kong, China

^f^ University of Malaya Medical Centre, Faculty of Medicine, Kuala Lumpur, Malaysia

^g^ Faculty of Law, University of Malaya, Kuala Lumpur Malaysia

^h^ Medical Research Ethics Committee, University of Malaya Medical Centre, Kuala Lumpur, Malaysia

^i^ Saw Swee Hock School of Public Health, National University of Singapore and National University Health System, Singapore, Singapore

^j^ London School of Hygiene & Tropical Medicine, London, England, United Kingdom

### Participant sampling and recruitment

In Singapore, survey participants were recruited through the Singapore Population Health Improvement Centre (SPHERiC) Online Panel, managed through the National University Health System to facilitate the dissemination of online surveys on wide-ranging topics concerning public health. The online panel comprises 2,500 English-literate Singaporeans or permanent residents who are aged 21 years and above. Members of the online panel were invited to take part in the survey via email or SMS sent on December 11, 2020. The invitation contained information regarding the purpose of the survey and a link to the questionnaire. The survey was administered in English through a web-based using the REDCap platform and took approximately 10 minutes to complete. A SGD$5 reimbursement was provided for completing the survey. Participants were given 10 days to complete the survey. A reminder was sent to participants who had not completed the e-questionnaire within 5 days. Data were anonymized prior to analysis.

In Malaysia, a market surveyor conducted the survey in English. Survey participants were Malaysians aged 18 years old and above, and purposively sampled to represent a mix of gender, ethnicities and locality (urban and suburban). Participant recruitment was conducted at public spaces such as shopping malls, community town-halls and residential halls with strict adherence to the standard operating procedure for safe distancing. The data collection was completed within five days (from 11th to 15th December 2020). All participants provided electronic consent and each participant took between 10 to 15 minutes to answer the questionnaire via an online link. No identifiable information was captured during the survey.

In Hong Kong, survey participants were recruited through the probability-based panel of the Hong Kong Public Opinion Research Institute. The panel includes 8,500 Cantonese- or English-literate Hong Kong citizens aged 18 years or above who are representative of specific sectors of the Hong Kong population. The survey invitations were sent to half of the members of the probability-based panel via email on November 30, 2020 and the other half of the members on December 8, 2020. Reminders were sent to those who had not completed the questionnaire by December 10, 2020. The invitation contained information regarding the purpose of the survey and a link to the e-questionnaire. The online platform was open until December 20, 2020. All questionnaires were self-administered by the respondents and submitted anonymously.

#### Tables S1-S3: Demographics of survey sample by territory

**Table S1**

Hong Kong: Comparison of survey sample with 2010 census population

|  |  | **Sample** | | **Census** |
| --- | --- | --- | --- | --- |
| **Variable** | **Groups** | **N (679)** | **%** | **%** |
| Age Group | 20 - 29 | 91 | 13.4 | 15.5 |
|  | 30 - 39 | 134 | 19.7 | 18.6 |
|  | 40 - 49 | 129 | 19.0 | 18.4 |
|  | 50 - 59 | 177 | 26.1 | 20.5 |
|  | 60 - 69 | 116 | 17.1 | 14.5 |
|  | 70+ | 32 | 4.7 | 12.4 |
| Gender | Female | 242 | 35.9 | 54.0 |
|  | Male | 432 | 64.1 | 46.0 |
| Ethnicity | Chinese | 621 | 91.9 | 92.0 |
|  | Indian |  | NA | 0.5 |
|  | Others | 55 | 8.1 | 7.5 |
| Marital Status | Married | 372 | 57.3 | 51.8 |
|  | Never married | 222 | 34.2 | 38.0 |
|  | Divorced | 36 | 5.5 | 4.1 |
|  | Separated | 7 | 1.1 | 0.4 |
|  | Widowed | 12 | 1.8 | 5.7 |
| Education Level | No formal qualifications/lower primary |  | NA | 12.0 |
|  | Primary | 2 | 0.3 | 13.7 |
|  | Junior secondary | 25 | 3.8 | 17.6 |
|  | Senior secondary | 128 | 19.2 | 26.1 |
|  | Tertiary, non-degree | 106 | 15.9 | 9.9 |
|  | Tertiary, degree | 404 | 60.8 | 20.7 |
| Monthly Household Income | Less than $10 000 | 36 | 6.3 | 19.1 |
|  | $10 000 to $19 999 | 64 | 11.3 | 21.8 |
|  | $20 000 to $29 999 | 84 | 14.8 | 15.8 |
|  | $30 000 or more | 383 | 67.5 | 43.2 |
| Housing | Public housing | 244 | 38.9 | 29.1 |
|  | Private housing | 370 | 59.0 | 53.2 |
|  | Others | 13 | 2.1 | 17.8 |

**Table S2**

Malaysia: Comparison of survey sample with 2010 census population

|  |  | **Sample** | | **Census** |
| --- | --- | --- | --- | --- |
| **Variable** | **Groups** | **N (1974)** | **%** | **%** |
| Age Group | 20 - 29 | 507 | 25.7 | 29.8 |
|  | 30 - 39 | 889 | 45.0 | 22.2 |
|  | 40 - 49 | 493 | 25.0 | 19.6 |
|  | 50 - 59 | 73 | 3.7 | 14.6 |
|  | 60 - 69 | 12 | 0.6 | 8.3 |
|  | 70+ |  | NA | 5.4 |
| Gender | Female | 1095 | 55.6 | 49.3 |
|  | Male | 876 | 44.4 | 50.7 |
| Ethnicity | Chinese | 993 | 50.4 | 24.5 |
|  | Malay | 641 | 32.5 | 67.3 |
|  | Indian | 334 | 17.0 | 7.3 |
|  | Others | 2 | 0.1 | 0.9 |
| Marital Status | Married | 886 | 46.4 | 59.6 |
|  | Single | 906 | 47.5 | 35.1 |
|  | Divorced/Separated | 85 | 4.5 | 0.8 |
|  | Widowed | 31 | 1.6 | 4.5 |
| Education Level | Primary | 8 | 0.4 | 17.5 |
|  | Junior secondary | 102 | 5.4 | 14.8 |
|  | Senior secondary | 818 | 43.0 | 41.5 |
|  | Tertiary, non-degree | 669 | 35.2 | 16.5 |
|  | Tertiary, degree | 305 | 16.0 | 9.7 |
| Monthly Household Income* | < RM4850 | 834 | 48.5 | 40.0 |
|  | RM4850 - 10959 | 715 | 41.5 | 40.0 |
|  | > RM 10959 | 172 | 10.0 | 20.0 |
| Housing | Flat/Apartment | 480 | 29.5 | 16.0 |
|  | Condominium/SOHO | 594 | 36.6 | 4.5 |
|  | Terrace/Townhouse | 467 | 28.7 | 35.4 |
|  | Semi-detached (Semi-D) | 68 | 4.2 | 7.1 |
|  | Bungalow/Penthouse | 14 | 0.9 | 34.4 |
|  | Others | 2 | 0.1 | 2.5 |

* Household Income & Basic Amenities Survey 2019 data was used for the "Census"

**Table S3**

Singapore: Comparison of survey sample with 2016 census population

|  |  | **Sample** | | **Census** |
| --- | --- | --- | --- | --- |
| **Variable** | **Groups** | **N (982)** | **%** | **%** |
| Age Group | 20 - 29 | 178 | 18.1 | 18.2 |
|  | 30 - 39 | 206 | 21.0 | 21.7 |
|  | 40 - 49 | 229 | 23.3 | 22.2 |
|  | 50 - 59 | 181 | 18.4 | 19.3 |
|  | 60 - 69 | 153 | 15.6 | 10.6 |
|  | 70+ | 35 | 3.6 | 8.0 |
| Gender | Female | 611 | 62.2 | 50.7 |
|  | Male | 371 | 37.8 | 49.3 |
| Ethnicity | Chinese | 821 | 83.6 | 74.1 |
|  | Malay | 46 | 4.7 | 13.4 |
|  | Indian | 91 | 9.3 | 9.2 |
|  | Others | 24 | 2.4 | 3.3 |
| Marital Status | Married | 571 | 58.6 | 59.4 |
|  | Single | 336 | 34.5 | 32.2 |
|  | Divorced/Separated | 55 | 5.6 | 3.3 |
|  | Widowed | 12 | 1.2 | 5.1 |
| Education Level | Below Secondary | 12 | 1.2 | 32.4 |
|  | Secondary | 160 | 16.4 | 18.9 |
|  | Post-Secondary, Non-University | 338 | 34.6 | 25.9 |
|  | University | 468 | 47.9 | 22.8 |
| Monthly Household Income | Less than $2000 | 143 | 16.0 | 11.8 |
|  | $2000 to $3999 | 179 | 20.1 | 18.4 |
|  | $4000 to $5999 | 175 | 19.6 | 17.1 |
|  | $6000 to $10000 | 248 | 27.8 | 24.2 |
|  | More than $10 000 | 147 | 16.5 | 28.5 |
| Housing | 1 to 2-room HDB | 29 | 3.0 | 4.6 |
|  | 3 - room HDB | 201 | 20.6 | 20.1 |
|  | 4 - room HDB | 370 | 37.9 | 32.0 |
|  | 5 - room HDB/executive flat | 269 | 27.6 | 25.7 |
|  | Condominium | 65 | 6.7 | 11.5 |
|  | Landed house | 32 | 3.3 | 5.7 |
|  | Others | 9 | 0.9 | 0.5 |

### Tables S4-S8: Numeric Data for Figures 1-5

#### Table S4

Numeric Data for Figure 1: Vaccine confidence and willingness to be vaccinated against COVID-19

| **Question** | **Response** | **Hong Kong** | **Malaysia** | **Singapore** |
| --- | --- | --- | --- | --- |
| Overall, I think vaccines are safe | Strongly Disagree | 30 (4%) | 22 (1%) | 40 (4%) |
|  | Disagree | 97 (14%) | 173 (9%) | 67 (7%) |
|  | Neither Agree nor Disagree | 266 (39%) | 449 (23%) | 321 (33%) |
|  | Agree | 248 (37%) | 821 (42%) | 393 (40%) |
|  | Strongly Agree | 37 (5%) | 509 (26%) | 161 (16%) |
| Overall, I think vaccines are effective | Strongly Disagree | 23 (3%) | 26 (1%) | 28 (3%) |
|  | Disagree | 57 (8%) | 174 (9%) | 55 (6%) |
|  | Neither Agree nor Disagree | 262 (39%) | 581 (29%) | 305 (31%) |
|  | Agree | 291 (43%) | 850 (43%) | 440 (45%) |
|  | Strongly Agree | 46 (7%) | 343 (17%) | 154 (16%) |
| Overall, I think vaccines are an important health intervention for children | Strongly Disagree | 28 (4%) | 27 (1%) | 32 (3%) |
|  | Disagree | 66 (10%) | 82 (4%) | 57 (6%) |
|  | Neither Agree nor Disagree | 236 (35%) | 461 (23%) | 237 (24%) |
|  | Agree | 268 (40%) | 794 (40%) | 379 (39%) |
|  | Strongly Agree | 80 (12%) | 610 (31%) | 277 (28%) |
| Vaccines are compatible with my religious beliefs | Strongly Disagree | 18 (5%) | 39 (2%) | 56 (7%) |
|  | Disagree | 24 (7%) | 70 (4%) | 41 (5%) |
|  | Neither Agree nor Disagree | 160 (45%) | 295 (15%) | 195 (25%) |
|  | Agree | 112 (31%) | 720 (36%) | 231 (30%) |
|  | Strongly Agree | 43 (12%) | 850 (43%) | 243 (32%) |
| I would be willing to get vaccinated against COVID-19 when an approved vaccine becomes available | Strongly Disagree | 26 (4%) | 32 (2%) | 68 (7%) |
|  | Disagree | 74 (11%) | 275 (14%) | 68 (7%) |
|  | Neither Agree nor Disagree | 190 (28%) | 569 (29%) | 233 (24%) |
|  | Agree | 302 (45%) | 600 (30%) | 320 (33%) |
|  | Strongly Agree | 86 (13%) | 498 (25%) | 293 (30%) |

#### Table S5

Numeric Data for Figure 2: Perceived intrusiveness of different control measures

| **Question** | **Response** | **Hong Kong** | **Malaysia** | **Singapore** |
| --- | --- | --- | --- | --- |
| Mandatory use of digital contact tracing technologies is an unreasonable restriction of my personal freedom | Strongly Disagree | 24 (4%) | 165 (8%) | 212 (22%) |
|  | Disagree | 70 (10%) | 184 (9%) | 287 (29%) |
|  | Neither Agree nor Disagree | 89 (13%) | 536 (27%) | 318 (32%) |
|  | Agree | 195 (29%) | 795 (40%) | 108 (11%) |
|  | Strongly Agree | 298 (44%) | 294 (15%) | 56 (6%) |
| Mandatory use of monitoring devices during quarantine is an unreasonable restriction of my personal freedom | Strongly Disagree | 41 (6%) | 165 (8%) | 255 (26%) |
|  | Disagree | 108 (16%) | 228 (12%) | 324 (33%) |
|  | Neither Agree nor Disagree | 154 (23%) | 522 (26%) | 259 (26%) |
|  | Agree | 209 (31%) | 775 (39%) | 86 (9%) |
|  | Strongly Agree | 164 (24%) | 284 (14%) | 58 (6%) |
| Requiring me to be vaccinated against COVID-19 before travelling would be an unreasonable infringement of my personal freedom | Strongly Disagree | 52 (8%) | 193 (10%) | 227 (23%) |
|  | Disagree | 220 (32%) | 169 (9%) | 312 (32%) |
|  | Neither Agree nor Disagree | 185 (27%) | 565 (29%) | 240 (24%) |
|  | Agree | 143 (21%) | 797 (40%) | 116 (12%) |
|  | Strongly Agree | 78 (12%) | 250 (13%) | 87 (9%) |
| Requiring me to show proof of a negative COVID-19 test before travelling is an unreasonable invasion of my privacy | Strongly Disagree | 98 (14%) | 224 (11%) | 372 (38%) |
|  | Disagree | 322 (48%) | 162 (8%) | 306 (31%) |
|  | Neither Agree nor Disagree | 163 (24%) | 494 (25%) | 188 (19%) |
|  | Agree | 54 (8%) | 837 (42%) | 70 (7%) |
|  | Strongly Agree | 39 (6%) | 257 (13%) | 46 (5%) |

#### Table S6

Numeric Data for Figure 3: Digital contact tracing: privacy, trust, and mandatory use

| **Question** | **Response** | **Hong Kong** | **Malaysia** | **Singapore** |
| --- | --- | --- | --- | --- |
| I trust that there are strict rules in my country/territory to prevent my digital contact tracing data from being used for other purposes | Strongly Disagree | 272 (40%) | 41 (2%) | 30 (3%) |
|  | Disagree | 192 (28%) | 138 (7%) | 57 (6%) |
|  | Neither Agree nor Disagree | 100 (15%) | 477 (24%) | 183 (19%) |
|  | Agree | 74 (11%) | 922 (47%) | 362 (37%) |
|  | Strongly Agree | 37 (5%) | 396 (20%) | 349 (36%) |
| I trust that there are strict rules in other countries/territories to prevent my digital contact tracing data from being used for other purposes when I travel there | Strongly Disagree | 66 (10%) | 41 (2%) | 114 (12%) |
|  | Disagree | 102 (15%) | 135 (7%) | 194 (20%) |
|  | Neither Agree nor Disagree | 250 (37%) | 504 (26%) | 384 (39%) |
|  | Agree | 194 (29%) | 894 (45%) | 166 (17%) |
|  | Strongly Agree | 63 (9%) | 400 (20%) | 123 (13%) |
| The benefits of digital contact tracing technologies outweigh their risks and burdens | Strongly Disagree | 130 (19%) | 1 (0%) | 22 (2%) |
|  | Disagree | 196 (29%) | 87 (4%) | 46 (5%) |
|  | Neither Agree nor Disagree | 198 (29%) | 505 (26%) | 275 (28%) |
|  | Agree | 111 (16%) | 994 (50%) | 411 (42%) |
|  | Strongly Agree | 41 (6%) | 387 (20%) | 227 (23%) |
| The use of digital contact tracing technologies should be mandatory during the COVID-19 pandemic | Strongly Disagree | 237 (35%) | 1 (0%) | 26 (3%) |
|  | Disagree | 166 (25%) | 70 (4%) | 28 (3%) |
|  | Neither Agree nor Disagree | 135 (20%) | 493 (25%) | 159 (16%) |
|  | Agree | 94 (14%) | 924 (47%) | 359 (37%) |
|  | Strongly Agree | 42 (6%) | 486 (25%) | 409 (42%) |
| Digital contact tracing helps to reduce the risk I might pose to others if I were to have a COVID-19 infection | Strongly Disagree | 89 (13%) | 5 (0%) | 19 (2%) |
|  | Disagree | 170 (25%) | 80 (4%) | 37 (4%) |
|  | Neither Agree nor Disagree | 153 (23%) | 509 (26%) | 159 (16%) |
|  | Agree | 209 (31%) | 958 (49%) | 425 (43%) |
|  | Strongly Agree | 52 (8%) | 422 (21%) | 341 (35%) |
| By using digital contact tracing technologies, I feel I can make a contribution to pandemic control efforts | Strongly Disagree | 145 (21%) | 11 (1%) | 15 (2%) |
|  | Disagree | 188 (28%) | 87 (4%) | 35 (4%) |
|  | Neither Agree nor Disagree | 182 (27%) | 553 (28%) | 177 (18%) |
|  | Agree | 123 (18%) | 937 (47%) | 432 (44%) |
|  | Strongly Agree | 40 (6%) | 386 (20%) | 322 (33%) |

#### Table S7

Numeric Data for Figure 4: Perceptions of COVID-19 vaccination for travel

| **Question** | **Response** | **Hong Kong** | **Malaysia** | **Singapore** |
| --- | --- | --- | --- | --- |
| It is reasonable to allow travel only for vaccinated people | Strongly Disagree | 39 (12%) | 30 (3%) | 44 (9%) |
|  | Disagree | 85 (25%) | 136 (14%) | 61 (12%) |
|  | Neither Agree nor Disagree | 71 (21%) | 138 (14%) | 122 (25%) |
|  | Agree | 113 (34%) | 360 (37%) | 167 (34%) |
|  | Strongly Agree | 26 (8%) | 315 (32%) | 97 (20%) |
| It is reasonable to require all international travelers to be vaccinated against COVID-19, unless there is a medical reason why they cannot be vaccinated | Strongly Disagree | 39 (12%) | 6 (1%) | 25 (5%) |
|  | Disagree | 70 (21%) | 125 (13%) | 38 (8%) |
|  | Neither Agree nor Disagree | 75 (23%) | 186 (19%) | 114 (23%) |
|  | Agree | 117 (35%) | 431 (44%) | 176 (36%) |
|  | Strongly Agree | 32 (10%) | 231 (24%) | 138 (28%) |
| Vaccinated individuals should be allowed to travel for any reason, while non-vaccinated individuals should only be allowed to travel for essential purposes (e.g. medical reasons, family emergencies, work) | Strongly Disagree | 56 (17%) | 8 (1%) | 35 (7%) |
|  | Disagree | 82 (25%) | 108 (11%) | 55 (11%) |
|  | Neither Agree nor Disagree | 82 (25%) | 212 (22%) | 130 (26%) |
|  | Agree | 87 (26%) | 389 (40%) | 181 (37%) |
|  | Strongly Agree | 26 (8%) | 262 (27%) | 90 (18%) |
| I would be willing to get a COVID-19 vaccine to travel abroad | Strongly Disagree | 29 (9%) | 7 (1%) | 23 (5%) |
|  | Disagree | 54 (16%) | 94 (10%) | 34 (7%) |
|  | Neither Agree nor Disagree | 79 (24%) | 220 (22%) | 90 (18%) |
|  | Agree | 133 (40%) | 339 (35%) | 202 (41%) |
|  | Strongly Agree | 39 (12%) | 319 (33%) | 142 (29%) |

#### Table S8

Numeric Data for Figure 5: Equity considerations in travel-related COVID-19 vaccination

| **Question** | **Response** | **Hong Kong** | **Malaysia** | **Singapore** |
| --- | --- | --- | --- | --- |
| Banning unvaccinated people from travelling internationally would be unfair | Strongly Disagree | 21 (6%) | 100 (10%) | 56 (11%) |
|  | Disagree | 60 (18%) | 113 (12%) | 78 (16%) |
|  | Neither Agree nor Disagree | 90 (27%) | 253 (26%) | 146 (30%) |
|  | Agree | 93 (28%) | 318 (32%) | 129 (26%) |
|  | Strongly Agree | 69 (21%) | 195 (20%) | 82 (17%) |
| It is reasonable to place different restrictions on vaccinated and unvaccinated travelers, if they pose different risks | Strongly Disagree | 14 (4%) | 11 (1%) | 21 (4%) |
|  | Disagree | 46 (14%) | 111 (11%) | 28 (6%) |
|  | Neither Agree nor Disagree | 72 (22%) | 207 (21%) | 114 (23%) |
|  | Agree | 162 (49%) | 382 (39%) | 208 (42%) |
|  | Strongly Agree | 40 (12%) | 268 (27%) | 120 (24%) |
| It is reasonable to require travelers to get vaccinated at their own expense, even if some groups in the population may not be able to afford to travel | Strongly Disagree | 16 (5%) | 85 (9%) | 43 (9%) |
|  | Disagree | 37 (11%) | 136 (14%) | 57 (12%) |
|  | Neither Agree nor Disagree | 93 (28%) | 247 (25%) | 161 (33%) |
|  | Agree | 153 (46%) | 325 (33%) | 167 (34%) |
|  | Strongly Agree | 34 (10%) | 186 (19%) | 63 (13%) |

### Tables S9-S11: Variables used to generate scores for vaccine confidence, support for use of monitoring devices during quarantine, and support for digital contact tracing

#### Table S9

The score for "Vaccine Confidence" is the mean of the likert scale values for the following 3 Likert scale questions

| **Response** | **Strongly Disagree** | **Disagree** | **Neither Agree nor Disagree** | **Agree** | **Strongly Agree** |
| --- | --- | --- | --- | --- | --- |
| **Score** | **1** | **2** | **3** | **4** | **5** |
| **Hong Kong** | | | | | |
| Overall, I think vaccines are safe | 30 (4%) | 97 (14%) | 266 (39%) | 248 (37%) | 37 (5%) |
| Overall, I think vaccines are effective | 23 (3%) | 57 (8%) | 262 (39%) | 291 (43%) | 46 (7%) |
| Overall, I think vaccines are an important health intervention for children | 28 (4%) | 66 (10%) | 236 (35%) | 268 (40%) | 80 (12%) |
| **Malaysia** | | | | | |
| Overall, I think vaccines are safe | 22 (1%) | 173 (9%) | 449 (23%) | 821 (42%) | 509 (26%) |
| Overall, I think vaccines are effective | 26 (1%) | 174 (9%) | 581 (29%) | 850 (43%) | 343 (17%) |
| Overall, I think vaccines are an important health intervention for children | 27 (1%) | 82 (4%) | 461 (23%) | 794 (40%) | 610 (31%) |
| **Singapore** | | | | | |
| Overall, I think vaccines are safe | 40 (4%) | 67 (7%) | 321 (33%) | 393 (40%) | 161 (16%) |
| Overall, I think vaccines are effective | 28 (3%) | 55 (6%) | 305 (31%) | 440 (45%) | 154 (16%) |
| Overall, I think vaccines are an important health intervention for children | 32 (3%) | 57 (6%) | 237 (24%) | 379 (39%) | 277 (28%) |

#### Table S10

The score for "Support for Wearable Monitoring Devices" is the mean of the likert scale values for the following 3 Likert scale questions

| **Response** | **Strongly Disagree** | **Disagree** | **Neither Agree nor Disagree** | **Agree** | **Strongly Agree** |
| --- | --- | --- | --- | --- | --- |
| **Score** | **1** | **2** | **3** | **4** | **5** |
| **Hong Kong** | | | | | |
| It is reasonable to require incoming travelers to my country/territory placed under quarantine to wear a monitoring device, even if they have not been infected or exposed to an infected person | 50 (7%) | 114 (17%) | 154 (23%) | 266 (39%) | 94 (14%) |
| Use of monitoring devices is an effective way of ensuring that people comply with quarantine orders | 69 (10%) | 139 (21%) | 163 (24%) | 236 (35%) | 71 (10%) |
| Mandatory use of monitoring devices during quarantine is necessary to limit risk to wider community | 96 (14%) | 124 (18%) | 168 (25%) | 206 (31%) | 81 (12%) |
| **Malaysia** | | | | | |
| It is reasonable to require incoming travelers to my country/territory placed under quarantine to wear a monitoring device, even if they have not been infected or exposed to an infected person | 4 (0%) | 87 (4%) | 316 (16%) | 890 (45%) | 677 (34%) |
| Use of monitoring devices is an effective way of ensuring that people comply with quarantine orders | 4 (0%) | 86 (4%) | 490 (25%) | 1049 (53%) | 345 (17%) |
| Mandatory use of monitoring devices during quarantine is necessary to limit risk to wider community | 10 (1%) | 124 (6%) | 551 (28%) | 888 (45%) | 401 (20%) |
| **Singapore** | | | | | |
| It is reasonable to require incoming travelers to my country/territory placed under quarantine to wear a monitoring device, even if they have not been infected or exposed to an infected person | 15 (2%) | 16 (2%) | 129 (13%) | 368 (37%) | 454 (46%) |
| Use of monitoring devices is an effective way of ensuring that people comply with quarantine orders | 13 (1%) | 20 (2%) | 100 (10%) | 366 (37%) | 483 (49%) |
| Mandatory use of monitoring devices during quarantine is necessary to limit risk to wider community | 16 (2%) | 36 (4%) | 133 (14%) | 384 (39%) | 413 (42%) |

#### Table S11

The score for "Support for Digital Contact Tracing" is the mean of the likert scale values for the following 6 Likert scale questions

| **Response** | **Strongly Disagree** | **Disagree** | **Neither Agree nor Disagree** | **Agree** | **Strongly Agree** |
| --- | --- | --- | --- | --- | --- |
| **Score** | **1** | **2** | **3** | **4** | **5** |
| **Hong Kong** | | | | | |
| Digital contact tracing technologies are effective for reducing the risk of COVID-19 spread | 80 (12%) | 185 (27%) | 170 (25%) | 192 (28%) | 49 (7%) |
| The benefits of digital contact tracing technologies outweigh their risks and burdens | 130 (19%) | 196 (29%) | 198 (29%) | 111 (16%) | 41 (6%) |
| Digital contact tracing helps to reduce my risk of COVID-19 | 148 (22%) | 195 (29%) | 180 (27%) | 120 (18%) | 33 (5%) |
| Digital contact tracing helps to reduce the risk I might pose to others if I were to have a COVID-19 infection | 89 (13%) | 170 (25%) | 153 (23%) | 209 (31%) | 52 (8%) |
| Digital contact tracing contributes to protecting public health and safety | 95 (14%) | 165 (24%) | 207 (30%) | 165 (24%) | 47 (7%) |
| Digital contact tracing protects vulnerable people who may get very sick from COVID-19 | 90 (13%) | 175 (26%) | 179 (26%) | 188 (28%) | 45 (7%) |
| **Malaysia** | | | | | |
| Digital contact tracing technologies are effective for reducing the risk of COVID-19 spread | 5 (0%) | 85 (4%) | 571 (29%) | 917 (46%) | 396 (20%) |
| The benefits of digital contact tracing technologies outweigh their risks and burdens | 1 (0%) | 87 (4%) | 505 (26%) | 994 (50%) | 387 (20%) |
| Digital contact tracing helps to reduce my risk of COVID-19 | 6 (0%) | 94 (5%) | 523 (26%) | 928 (47%) | 423 (21%) |
| Digital contact tracing helps to reduce the risk I might pose to others if I were to have a COVID-19 infection | 5 (0%) | 80 (4%) | 509 (26%) | 958 (49%) | 422 (21%) |
| Digital contact tracing contributes to protecting public health and safety | 3 (0%) | 91 (5%) | 487 (25%) | 1003 (51%) | 390 (20%) |
| Digital contact tracing protects vulnerable people who may get very sick from COVID-19 | 11 (1%) | 117 (6%) | 518 (26%) | 957 (48%) | 371 (19%) |
| **Singapore** | | | | | |
| Digital contact tracing technologies are effective for reducing the risk of COVID-19 spread | 18 (2%) | 47 (5%) | 181 (18%) | 424 (43%) | 311 (32%) |
| The benefits of digital contact tracing technologies outweigh their risks and burdens | 22 (2%) | 46 (5%) | 275 (28%) | 411 (42%) | 227 (23%) |
| Digital contact tracing helps to reduce my risk of COVID-19 | 46 (5%) | 82 (8%) | 208 (21%) | 386 (39%) | 259 (26%) |
| Digital contact tracing helps to reduce the risk I might pose to others if I were to have a COVID-19 infection | 19 (2%) | 37 (4%) | 159 (16%) | 425 (43%) | 341 (35%) |
| Digital contact tracing contributes to protecting public health and safety | 16 (2%) | 28 (3%) | 153 (16%) | 419 (43%) | 365 (37%) |
| Digital contact tracing protects vulnerable people who may get very sick from COVID-19 | 28 (3%) | 55 (6%) | 192 (20%) | 393 (40%) | 313 (32%) |

**Tables S12-S14: Willingness to be vaccinated against COVID-19 by territory**

#### Table S12

Hong Kong: Willingness to be vaccinated against COVID-19 by vaccine confidence score and demographic characteristics

|  | **Unwilling (N=100)** | **Undecided (N=190)** | **Willing (N=388)** | **Total (N=678)** | **p-value** |
| --- | --- | --- | --- | --- | --- |
| **Vaccine Confidence** |  |  |  |  | < 0.001 |
| 1 - 2 | 37 (63.8%) | 14 (24.1%) | 7 (12.1%) | 58 (100.0%) |  |
| > 2, up to 3 | 44 (18.6%) | 104 (44.1%) | 88 (37.3%) | 236 (100.0%) |  |
| > 3, up to 4 | 19 (6.1%) | 65 (20.8%) | 229 (73.2%) | 313 (100.0%) |  |
| > 4, up to 5 | 0 (0.0%) | 7 (9.9%) | 64 (90.1%) | 71 (100.0%) |  |
| **Age Group** |  |  |  |  | 0.174 |
| 18 - 29 | 18 (19.8%) | 19 (20.9%) | 54 (59.3%) | 91 (100.0%) |  |
| 30 - 39 | 16 (11.9%) | 40 (29.9%) | 78 (58.2%) | 134 (100.0%) |  |
| 40 - 49 | 15 (11.6%) | 34 (26.4%) | 80 (62.0%) | 129 (100.0%) |  |
| 50 - 59 | 30 (16.9%) | 57 (32.2%) | 90 (50.8%) | 177 (100.0%) |  |
| 60 - 69 | 20 (17.2%) | 33 (28.4%) | 63 (54.3%) | 116 (100.0%) |  |
| 70+ | 1 (3.2%) | 7 (22.6%) | 23 (74.2%) | 31 (100.0%) |  |
| **Gender** |  |  |  |  | < 0.001 |
| Female | 51 (21.1%) | 84 (34.7%) | 107 (44.2%) | 242 (100.0%) |  |
| Male | 48 (11.1%) | 106 (24.6%) | 277 (64.3%) | 431 (100.0%) |  |
| Non-binary | 0 (0.0%) | 0 (0.0%) | 3 (100.0%) | 3 (100.0%) |  |
| **Ethnicity** |  |  |  |  | 0.342 |
| Chinese | 94 (15.2%) | 171 (27.6%) | 355 (57.3%) | 620 (100.0%) |  |
| Others | 5 (9.1%) | 19 (34.5%) | 31 (56.4%) | 55 (100.0%) |  |
| **Marital Status** |  |  |  |  | 0.247 |
| Currently married | 50 (13.5%) | 100 (27.0%) | 221 (59.6%) | 371 (100.0%) |  |
| Never married | 40 (18.0%) | 59 (26.6%) | 123 (55.4%) | 222 (100.0%) |  |
| Others* | 5 (9.1%) | 20 (36.4%) | 30 (54.5%) | 55 (100.0%) |  |
| **Education Level** |  |  |  |  | 0.105 |
| Secondary or below | 17 (11.0%) | 52 (33.8%) | 85 (55.2%) | 154 (100.0%) |  |
| Tertiary, non-degree | 15 (14.2%) | 34 (32.1%) | 57 (53.8%) | 106 (100.0%) |  |
| Tertiary, degree | 66 (16.3%) | 97 (24.0%) | 241 (59.7%) | 404 (100.0%) |  |
| **Monthly Household Income** |  |  |  |  | 0.711 |
| Less than $10 000 | 6 (16.7%) | 13 (36.1%) | 17 (47.2%) | 36 (100.0%) |  |
| $10 000 to $19 999 | 11 (17.2%) | 18 (28.1%) | 35 (54.7%) | 64 (100.0%) |  |
| $20 000 to $29 999 | 12 (14.3%) | 28 (33.3%) | 44 (52.4%) | 84 (100.0%) |  |
| $30 000 to $50 000 | 20 (13.2%) | 43 (28.3%) | 89 (58.6%) | 152 (100.0%) |  |
| More than $50 000 | 30 (13.0%) | 57 (24.8%) | 143 (62.2%) | 230 (100.0%) |  |
| **Housing** |  |  |  |  | 0.732 |
| 1 to 2-room public housing | 28 (16.4%) | 43 (25.1%) | 100 (58.5%) | 171 (100.0%) |  |
| 3 - room public housing | 12 (17.1%) | 18 (25.7%) | 40 (57.1%) | 70 (100.0%) |  |
| Private housing estates | 48 (13.3%) | 107 (29.6%) | 206 (57.1%) | 361 (100.0%) |  |
| Others** | 4 (16.7%) | 4 (16.7%) | 16 (66.7%) | 24 (100.0%) |  |

* Marital Status - "Others" comprises of "Divorced", "Separated but not divorced" & "Widowed"

** Housing - "Others" comprises of "4 - room public housing", "5 - room public housing", "Landed house" & "Others"

Refuse to answer

Gender: n = 2

Ethnicity: n = 3

Marital Status: n = 30

Education Level: n = 14

Monthly Household Income: n = 95

Housing: n = 48

Do not know

Monthly Household Income: n = 17

Housing: n = 4

#### Table S13

Malaysia: Willingness to be vaccinated against COVID-19 by vaccine confidence score and demographic characteristics

|  | **Unwilling (N=307)** | **Undecided (N=569)** | **Willing (N=1098)** | **Total (N=1974)** | **p-value** |
| --- | --- | --- | --- | --- | --- |
| **Vaccine Confidence** |  |  |  |  | < 0.001 |
| 1 - 2 | 51 (87.9%) | 6 (10.3%) | 1 (1.7%) | 58 (100.0%) |  |
| > 2, up to 3 | 77 (29.5%) | 109 (41.8%) | 75 (28.7%) | 261 (100.0%) |  |
| > 3, up to 4 | 152 (14.9%) | 305 (30.0%) | 560 (55.1%) | 1017 (100.0%) |  |
| > 4, up to 5 | 27 (4.2%) | 149 (23.4%) | 462 (72.4%) | 638 (100.0%) |  |
| **Age Group** |  |  |  |  | < 0.001 |
| 18 - 29 | 58 (11.4%) | 134 (26.4%) | 315 (62.1%) | 507 (100.0%) |  |
| 30 - 39 | 134 (15.1%) | 240 (27.0%) | 515 (57.9%) | 889 (100.0%) |  |
| 40 - 49 | 99 (20.1%) | 168 (34.1%) | 226 (45.8%) | 493 (100.0%) |  |
| 50 - 59 | 13 (17.8%) | 23 (31.5%) | 37 (50.7%) | 73 (100.0%) |  |
| 60 - 69 | 3 (25.0%) | 4 (33.3%) | 5 (41.7%) | 12 (100.0%) |  |
| **Gender** |  |  |  |  | 0.009 |
| Female | 149 (13.6%) | 303 (27.7%) | 643 (58.7%) | 1095 (100.0%) |  |
| Male | 157 (17.9%) | 266 (30.4%) | 453 (51.7%) | 876 (100.0%) |  |
| Non-binary | 1 (50.0%) | 0 (0.0%) | 1 (50.0%) | 2 (100.0%) |  |
| **Ethnicity** |  |  |  |  | 0.482 |
| Chinese | 152 (15.3%) | 279 (28.1%) | 562 (56.6%) | 993 (100.0%) |  |
| Malay | 94 (14.7%) | 186 (29.0%) | 361 (56.3%) | 641 (100.0%) |  |
| Indian | 60 (18.0%) | 102 (30.5%) | 172 (51.5%) | 334 (100.0%) |  |
| Others | 1 (50.0%) | 0 (0.0%) | 1 (50.0%) | 2 (100.0%) |  |
| **Marital Status** |  |  |  |  | < 0.001 |
| Currently married | 152 (17.2%) | 285 (32.2%) | 449 (50.7%) | 886 (100.0%) |  |
| Never married | 106 (11.7%) | 227 (25.1%) | 573 (63.2%) | 906 (100.0%) |  |
| Others* | 35 (30.2%) | 39 (33.6%) | 42 (36.2%) | 116 (100.0%) |  |
| **Education Level** |  |  |  |  | 0.003 |
| Junior secondary or below | 22 (16.7%) | 32 (24.2%) | 78 (59.1%) | 132 (100.0%) |  |
| Senior secondary | 104 (12.7%) | 257 (31.4%) | 457 (55.9%) | 818 (100.0%) |  |
| Tertiary, non-degree | 102 (15.2%) | 188 (28.1%) | 379 (56.7%) | 669 (100.0%) |  |
| Tertiary, degree | 69 (22.6%) | 80 (26.2%) | 156 (51.1%) | 305 (100.0%) |  |
| **Monthly Household Income** |  |  |  |  | < 0.001 |
| < RM4850 | 83 (10.0%) | 223 (26.7%) | 528 (63.3%) | 834 (100.0%) |  |
| RM4850 - 10959 | 139 (19.4%) | 230 (32.2%) | 346 (48.4%) | 715 (100.0%) |  |
| > RM 10959 | 52 (30.2%) | 52 (30.2%) | 68 (39.5%) | 172 (100.0%) |  |
| **Housing** |  |  |  |  | 0.014 |
| Flat/Apartment | 58 (12.1%) | 138 (28.8%) | 284 (59.2%) | 480 (100.0%) |  |
| Condominium/SOHO | 112 (18.9%) | 185 (31.1%) | 297 (50.0%) | 594 (100.0%) |  |
| Terrace/Townhouse | 85 (18.2%) | 131 (28.1%) | 251 (53.7%) | 467 (100.0%) |  |
| Semi-detached (Semi-D) | 14 (20.6%) | 20 (29.4%) | 34 (50.0%) | 68 (100.0%) |  |
| Others** | 1 (6.2%) | 2 (12.5%) | 13 (81.2%) | 16 (100.0%) |  |

* Marital Status - "Others" comprises of "Divorced", "Separated but not divorced" & "Widowed"

** Housing - "Others" comprises of "Bungalow/Penthouse" & "Others"

Refuse to answer

Marital Status: n = 60

Education Level: n = 39

Monthly Household Income: n = 213

Housing: n = 192

Do not know

Monthly Household Income: n = 34

Housing: n = 154

NA

Gender: n = 1

Ethnicity: n = 4

Marital Status: n = 6

Education Level: n = 11

Monthly Household Income: n = 6

Housing: n = 3

#### Table S14

Singapore: Willingness to be vaccinated against COVID-19 by vaccine confidence score and demographic characteristics

|  | **Unwilling (N=136)** | **Undecided (N=233)** | **Willing (N=613)** | **Total (N=982)** | **p-value** |
| --- | --- | --- | --- | --- | --- |
| **Vaccine Confidence** |  |  |  |  | < 0.001 |
| 1 - 2 | 47 (78.3%) | 9 (15.0%) | 4 (6.7%) | 60 (100.0%) |  |
| > 2, up to 3 | 57 (27.0%) | 102 (48.3%) | 52 (24.6%) | 211 (100.0%) |  |
| > 3, up to 4 | 22 (4.9%) | 100 (22.2%) | 328 (72.9%) | 450 (100.0%) |  |
| > 4, up to 5 | 10 (3.8%) | 22 (8.4%) | 229 (87.7%) | 261 (100.0%) |  |
| **Age Group** |  |  |  |  | < 0.001 |
| 18 - 29 | 15 (8.4%) | 33 (18.5%) | 130 (73.0%) | 178 (100.0%) |  |
| 30 - 39 | 26 (12.6%) | 42 (20.4%) | 138 (67.0%) | 206 (100.0%) |  |
| 40 - 49 | 28 (12.2%) | 51 (22.3%) | 150 (65.5%) | 229 (100.0%) |  |
| 50 - 59 | 28 (15.5%) | 54 (29.8%) | 99 (54.7%) | 181 (100.0%) |  |
| 60 - 69 | 34 (22.2%) | 40 (26.1%) | 79 (51.6%) | 153 (100.0%) |  |
| 70+ | 5 (14.3%) | 13 (37.1%) | 17 (48.6%) | 35 (100.0%) |  |
| **Gender** |  |  |  |  | < 0.001 |
| Female | 94 (15.4%) | 165 (27.0%) | 352 (57.6%) | 611 (100.0%) |  |
| Male | 42 (11.3%) | 68 (18.3%) | 261 (70.4%) | 371 (100.0%) |  |
| **Ethnicity** |  |  |  |  | 0.056 |
| Chinese | 120 (14.6%) | 197 (24.0%) | 504 (61.4%) | 821 (100.0%) |  |
| Malay | 1 (2.2%) | 12 (26.1%) | 33 (71.7%) | 46 (100.0%) |  |
| Indian | 11 (12.1%) | 15 (16.5%) | 65 (71.4%) | 91 (100.0%) |  |
| Others | 4 (16.7%) | 9 (37.5%) | 11 (45.8%) | 24 (100.0%) |  |
| **Marital Status** |  |  |  |  | 0.495 |
| Currently married | 71 (12.4%) | 134 (23.5%) | 366 (64.1%) | 571 (100.0%) |  |
| Never married | 49 (14.6%) | 80 (23.8%) | 207 (61.6%) | 336 (100.0%) |  |
| Others* | 13 (19.4%) | 17 (25.4%) | 37 (55.2%) | 67 (100.0%) |  |
| **Education Level** |  |  |  |  | 0.028 |
| Secondary or below | 21 (12.2%) | 52 (30.2%) | 99 (57.6%) | 172 (100.0%) |  |
| Post-Secondary, Non-University | 46 (13.6%) | 90 (26.6%) | 202 (59.8%) | 338 (100.0%) |  |
| University | 68 (14.5%) | 90 (19.2%) | 310 (66.2%) | 468 (100.0%) |  |
| **Monthly Household Income** |  |  |  |  | 0.002 |
| Less than $2000 | 31 (21.7%) | 46 (32.2%) | 66 (46.2%) | 143 (100.0%) |  |
| $2000 to $3999 | 24 (13.4%) | 45 (25.1%) | 110 (61.5%) | 179 (100.0%) |  |
| $4000 to $5999 | 25 (14.3%) | 35 (20.0%) | 115 (65.7%) | 175 (100.0%) |  |
| $6000 to $10000 | 29 (11.7%) | 54 (21.8%) | 165 (66.5%) | 248 (100.0%) |  |
| More than $10 000 | 17 (11.6%) | 26 (17.7%) | 104 (70.7%) | 147 (100.0%) |  |
| **Housing** |  |  |  |  | 0.193 |
| 3 - room HDB | 26 (12.9%) | 56 (27.9%) | 119 (59.2%) | 201 (100.0%) |  |
| 4 - room HDB | 41 (11.1%) | 87 (23.5%) | 242 (65.4%) | 370 (100.0%) |  |
| 5 - room HDB/executive flat | 43 (16.0%) | 56 (20.8%) | 170 (63.2%) | 269 (100.0%) |  |
| Condominium | 15 (23.1%) | 15 (23.1%) | 35 (53.8%) | 65 (100.0%) |  |
| Others** | 10 (14.3%) | 17 (24.3%) | 43 (61.4%) | 70 (100.0%) |  |

* Marital Status - "Others" comprises of "Divorced", "Separated but not divorced" & "Widowed"

** Housing - "Others" comprises of "1 to 2-room HDB", "Landed house" & "Others"

Refuse to answer

Marital Status: n = 8

Education Level: n = 4

Monthly Household Income: n = 49

Housing: n = 5

Do not know

Monthly Household Income: n = 41

Housing: n = 2

### Figures S1-S2: Willingness to be vaccinated against COVID-19 and support for monitoring devices and digital contact tracing

#### Figure S1:

#### Percentage of respondents willing to be vaccinated against COVID-19, with Wilson score 95% confidence intervals (y-axis), level of support for digital contact tracing* (x-axis) and territory

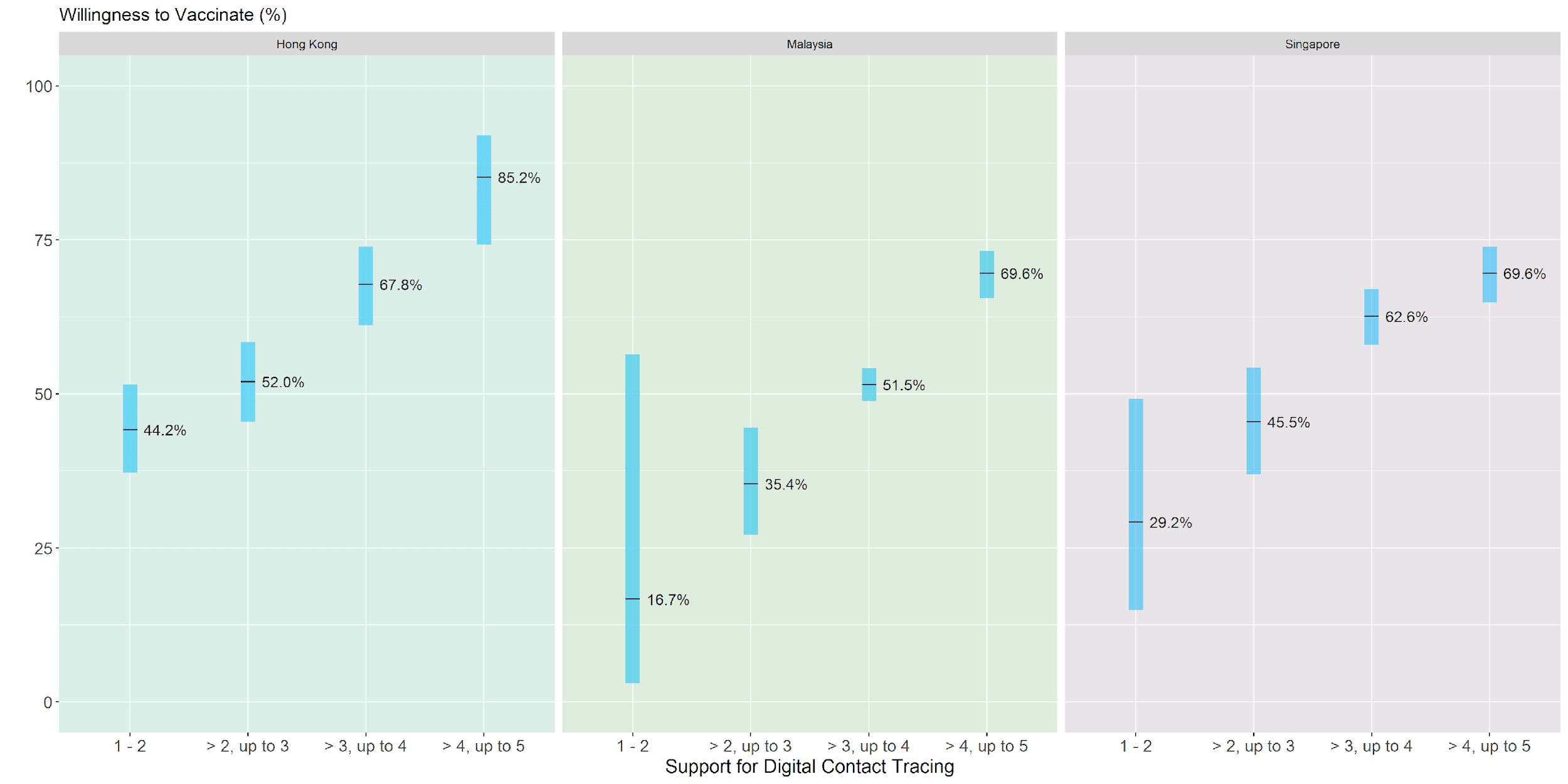

* The score for "Support for Digital Contact Tracing" is the mean of the likert scale values for the following 6 Likert scale questions:

- Digital contact tracing technologies are effective for reducing the risk of COVID-19 spread
- The benefits of digital contact tracing technologies outweigh their risks and burdens
- Digital contact tracing helps to reduce my risk of COVID-19
- Digital contact tracing helps to reduce the risk I might pose to others if I were to have a COVID-19 infection
- Digital contact tracing contributes to protecting public health and safety
- Digital contact tracing protects vulnerable people who may get very sick from COVID-19

#### Figure S2:

#### Percentage of respondents willing to be vaccinated against COVID-19, with Wilson score 95% confidence intervals (y-axis), level of support for digital contact tracing* (x-axis), and territory

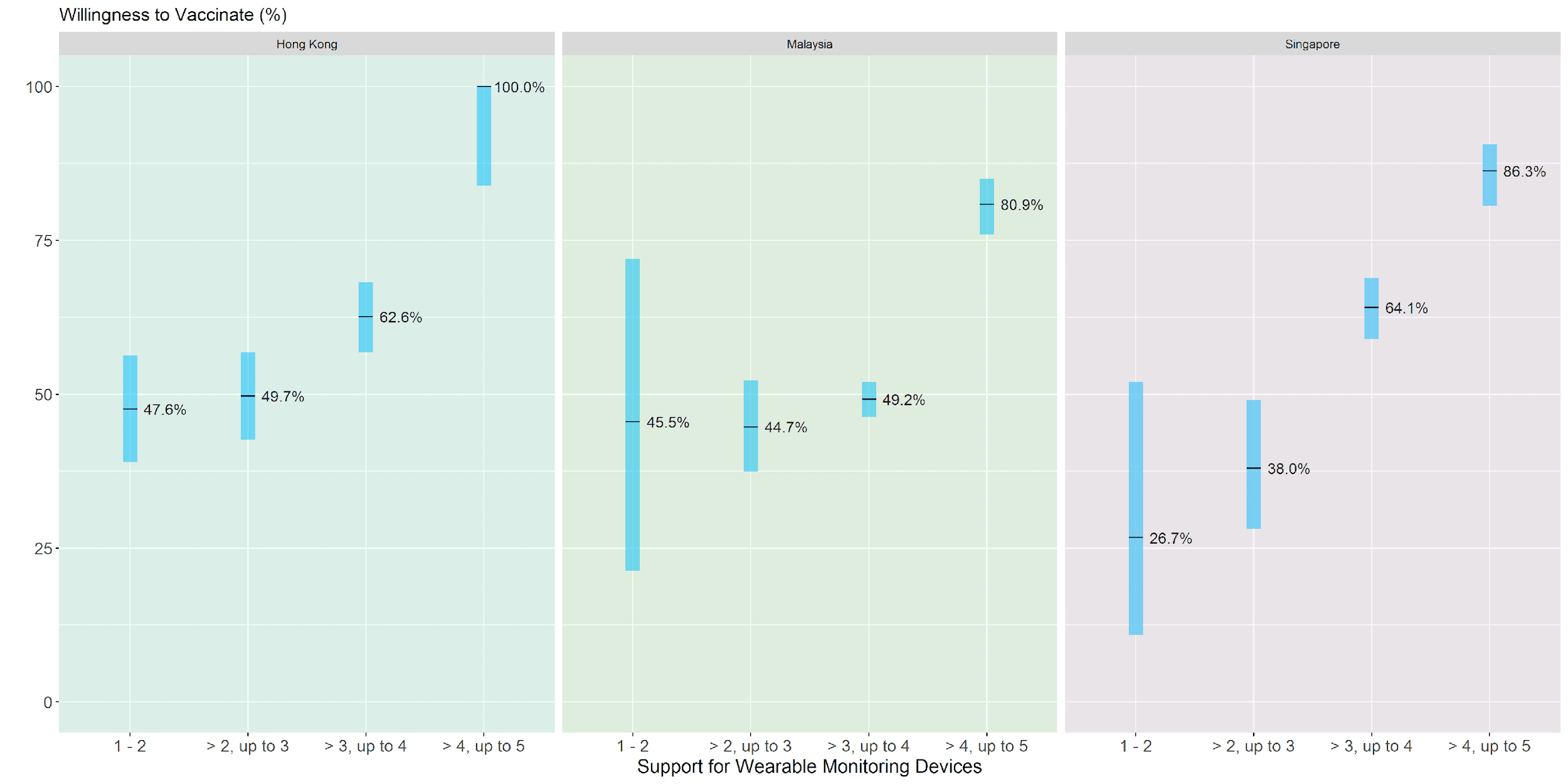

* The score for "Support for Wearable Monitoring Devices" is the mean of the likert scale values for the following 3 Likert scale questions:

- It is reasonable to require incoming travelers to my country/territory placed under quarantine to wear a monitoring device, even if they have not been infected or exposed to an infected person
- Use of monitoring devices is an effective way of ensuring that people comply with quarantine orders
- Mandatory use of monitoring devices during quarantine is necessary to limit risk to wider community
